# A pitfall in estimating the effective reproductive number *R_t_* for COVID-19

**DOI:** 10.1101/2020.05.12.20099366

**Authors:** Markus Petermann, Daniel Wyler

## Abstract

The effective reproductive number *R*_*t*_ of COVID-19 is determined indirectly from data that are only incompletely known. Approaches based on reconstructing these data by sampling time lags from suitable distributions introduce noise effects that can result in distorted estimates of *R*_*t*_. This, in turn, may lead to misleading interpretations of the efficacy of the various measures taken to limit COVID-19 transmission. We discuss in some detail a study used for real time monitoring of the reproductive number in Switzerland; see https://ncs-tf.ch/en/situation-report.

We argue that the method used to derive the above curve is systematically flawed and leads to an underestimation of the efficacy of the lockdown. The method adopted by the Robert Koch Institute suffers from similar deficiencies, their impact is however smaller.

## Introduction

The daily varying effective reproductive number *R*_*t*_ is often used to monitor the spread of epidemic diseases such as COVID-19. It measures the expected number of secondary infections on day *t* due to a single infected individual and is given by

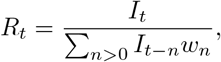

where *I*_*t*_ is the number of new infections on day *t* (and equally *I*_*t*−*n*_ the number of infections on day *t*−*n* and so on), and the *w*_*n*_ are the infection intensities, i.e. *w*_*n*_ is the probability that a secondary infection was contracted from a person who got infected *n* days earlier. While the infection intensities can be fitted to available data, the *I*_*t*_ are not observable directly, unless representative proportions of the population were tested on a daily basis. Therefore, they need to be inferred indirectly from some other data.

There exist different schemes to reconstruct the *I*_*t*_ from the data, like the classical statistical inference methods. In the present article we concentrate on schemes that are based on the idea of a ”mechanical” reconstruction of the infection data by sampling time lags of observed data from suitable distributions. In the context of the current COVID-19 pandemic, such schemes have been implemented in different ways by various groups in different countries.

In the following section, we present these schemes and show how they systematically introduce noise into the true data. In the remaining sections we examine the impact of the noise on the reproductive numbers calculated from these data.

### The reconstruction scheme and its smoothing effect

We exemplify the scheme and how it introduces noise into the data by the version implemented by Scire et al [2], that is very similar to the implementation by Abbott s, Hellewell J, Thompson RNN et al [3]. For the sake of clarity, we limit our exposition of the scheme to the observables *C*_*t*_, the number of confirmed cases on day *t*. The other observables used analogously by the group are hospitalizations and deaths. Moreover, as our focus is on the days around March 17, when the lockdown started, and as more than two months have passed since then, we restrict our exposition to those days of infection where all infections can be assumed to be confirmed by the actual date of the monitoring. According to the parameters used by the group, 95% of the cases are confirmed within 20 days after infection. For their method of extending the reconstruction to later days we refer again to their article.

Now, let *X* denote the incubation time of a randomly drawn case, i.e. the time between infection and symptom onset, and analogously *Y* the time between symptom onset and confirmation. The distributions of *X* and *Y* result from fitting to available data; see [2] and references therein. Then for every confirmed case *a* one samples independently a 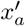 from the distribution of *X* and a 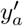 from the distribution of *Y*.

The *reconstructed* infection day 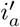 of this case *a* is then simply the day when the virus infection was confirmed minus 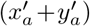. Counting the number of cases that fall now on day *t* gives the reconstructed *I*_*t*_, that we denote by 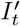. The reproductive numbers calculated from these 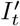 are denoted by 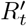.

That this scheme introduces noise into the true data is seen as follows. We denote the true infection day by *i*_*a*_, the true incubation period by *x*_*a*_ and the true time between symptom onset and confirmation by *y*_*a*_. Then we have 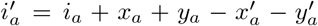. As the sampled 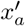 and 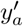 are independent of the true *x*_*a*_ and *y*_*a*_ (because we don’t know these; we just know that they are approximately distributed like *X* and *Y*, respectively), the reconstructed 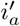 equals the true ”signal” *i*_*a*_ plus some ”noise” 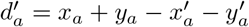.

As we will see in the now following examples, this results in a smoothing of the infection number statistics^1^, which in turn, under certain circumstances, has a significant impact on the reproductive numbers calculated from it.

### An illustrative example

The following example illustrates the effect of this scheme on the reconstructed reproductive number. Assume that *I*_1_ = 128 and *R*_*t*_ = 2 for *t* ≤ 6 and *R*_*t*_ = 0.8 for *t* ≥ 7, and that infectiousness is limited to the day after infection, i.e. *w*_1_ = 1. This yields the ’true’ infection numbers and reproductive numbers which are illustrated by the red curves in Fig. 2. For the *reconstructed* data we take *X* and *Y* both to be gaussian with mean 5 and standard deviation 1. Thus, the ”noise” is also gaussian with mean 0 and standard deviation 2. With this ”noise”, the scheme results in the average in the corresponding blue curves. Of course, the blue curve *R*′ is prone to lead to wrong decisions: If a lockdown caused the sharp decline in *R* from day 6 to day 7, then the blue curve may suggest that its impact was much less important and that most of the reduction was achieved already before the lockdown. It might even lead to the conclusion that the lockdown was not needed at all and that softer measures in force already before day 6 have had a sufficient effect, where in reality they had no effect at all. This in turn could lead to the conclusion, that the pandemic can be kept under control by adhering to soft measures only.

**Figure 1:**
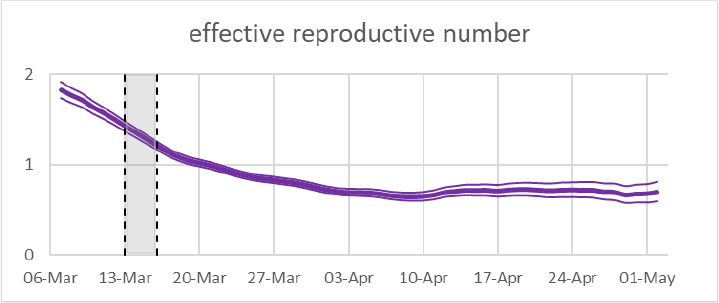
Effective reproductive numbers in Switzerland as shown in the real time monitoring on May 13. Mean and 95% uncertainty intervals are estimated on confirmed cases [1]. The shaded region is the onset of strong public health measures.

**Figure 2:**
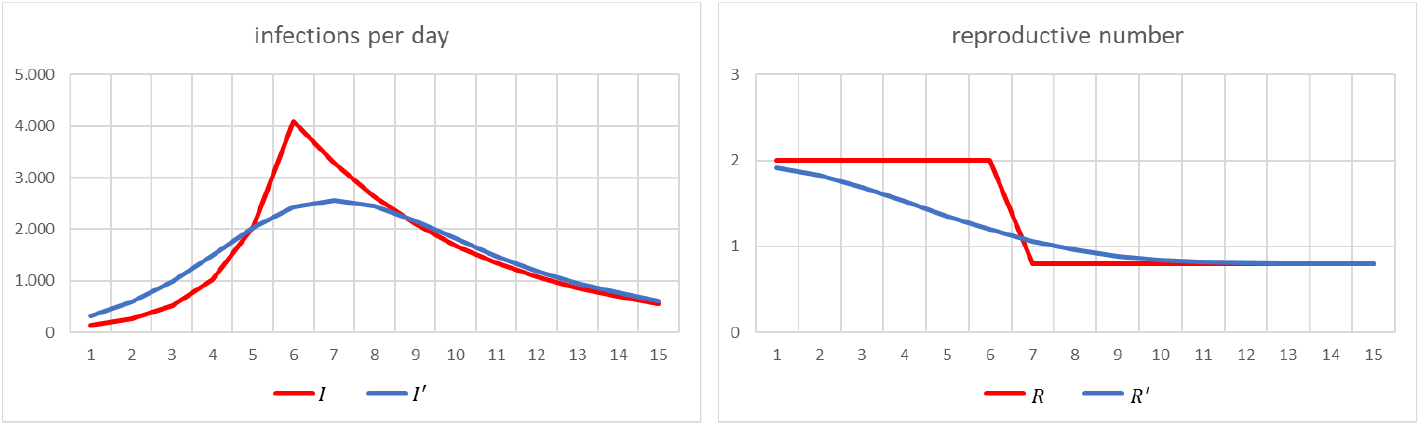
’True’ (red) and reconstructed (blue) infection and reproductive numbers

### The reproductive number in Switzerland around March 17

We now turn to the situation in Switzerland around March 17, 2020. A calculation based on distributions for the time lags *X* and *Y* and the infection intensity as described in [2] yields the following result:

Unlike in the illustrative example above, we cannot start from ’true’ numbers of new infections. We instead choose^2^ the numbers of new infections, denoted by *I*^*fit*^ (red curve), in such a way that the resulting expected numbers of confirmed cases *C*^*av*^ (solid black curve) fit well the black dots *C*^*true*^ which show the actually reported data of confirmed cases [4]. Here, to get *C*^*av*^ from the numbers of new infections, we shift forward the infection day of each such case by sampling independently from *X* and *Y*.

Given *I*^*fit*^, we proceed as in the illustrative example, but, of course, with ”noise” according to these *X* and *Y* instead of the gaussians used there. This gives the *reconstructed* numbers of new infections *I*′ (blue curve). Using the infection intensities *w*_*n*_ from [2], we calculate the corresponding reproductive numbers *R*^*fit*^ and *R*′. The latter matches well the green curve *R*^*TF*^ that shows the estimated mean reproductive numbers, as published on the website [1] on May 13.

The remarks made above on the illustrative example apply also here. We note also that our red curve of reproductive numbers is in quite good agreement with the results of the inference analysis reported in [5, 6].

### An alternative approach to reconstruction

Contrary to Switzerland, where, as far as we know, the date of symptom onset of the single cases is not collected systematically, this information is available for the majority of the cases in Germany. For the sake of clarity, we assume here that it is known for all cases.^3^ Then, the Robert Koch Institute applies the following simpler scheme [7]:^4^ The reconstructed infection day 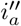 of a case *a* is the day of its symptom onset minus *m*, where *m* is the average incubation time. Counting the number of cases that fall now on day *t* gives the reconstructed *I*_*t*_ that we denote by 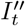. The reproductive numbers calculated from these 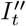 are denoted by 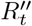.

Of course we can view the subtracted value *m* as sample from the distribution of the constant time lag *X* = *m*. Therefore, this scheme is at least formally very similar to the one adopted by [2].

From the above discussion it is now clear that the so reconstructed infection times 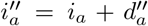 with ”noise” 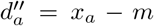 lead also to a smoothing of the infection number statistics and thus to misleading reproductive numbers. But it is also intuitively clear, that the impact is significantly smaller. This is confirmed by the following calculation.

Assume that the above *I*_*fit*_ are the true new infections per day, and that the distribution of the incubation time *X* is as above. Denote by 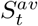 the resulting expected number of cases with symptom onset on day *t*. Then 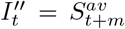. Assuming moreover that also the infection intensities are as above, we get the following result: We stress that the knowledge of the dates of symptom onset is an advantage, as adopting the same scheme but with dates of confirmation instead of symptom onset, would introduce the ”noise” *x*_*a*_ + *y*_*a*_ *–m*′, where *m*′ is the average time between infection and confirmation, into the true data, and this is clearly more ”noise” than in the scheme based on dates of symptom onset.

## Conclusion

In this note we have reexamined a type of schemes used to estimate the effective reproductive numbers *R*_*t*_ for COVID-19 by the example of two versions actually in use [2, 7]. These schemes are based on reconstruction of not directly observable data by sampling time lags of observed data from suitable distributions. Noise effects inherent in these schemes smooth the number statistics of the true data. The analysis of thus smoothed number statistics yields stable results and is easier to handle than classical inference methods as applied in [5, 6]. However, under certain circumstances like the current COVID-19 pandemic, the introduced noise effects dominate the information contained in the true data and lead to erroneous interpretations. The simpler approach adopted by [7] performs better than the one used in [2]. Moreover, we point out that adequate knowledge of the date of symptom onset is an advantage.

## Data Availability

https://github.com/daenuprobst/covid19-cases-switzerland
https://bsse.ethz.ch/cevo/research/sars-cov-
2/real-time-monitoring-in-switzerland.html.

## Appendix A Fitting algorithm

Our algorithm leading to the curves *I*^*fit*^ and *R*^*fit*^ of Fig. 3 can be viewed as a deterministic or non-Bayesian version of the analysis performed in [6] to estimate the effects of non-pharmaceutical interventions on COVID-19 in Europe. We give in the following the details of our algorithm.

**Figure 3:**
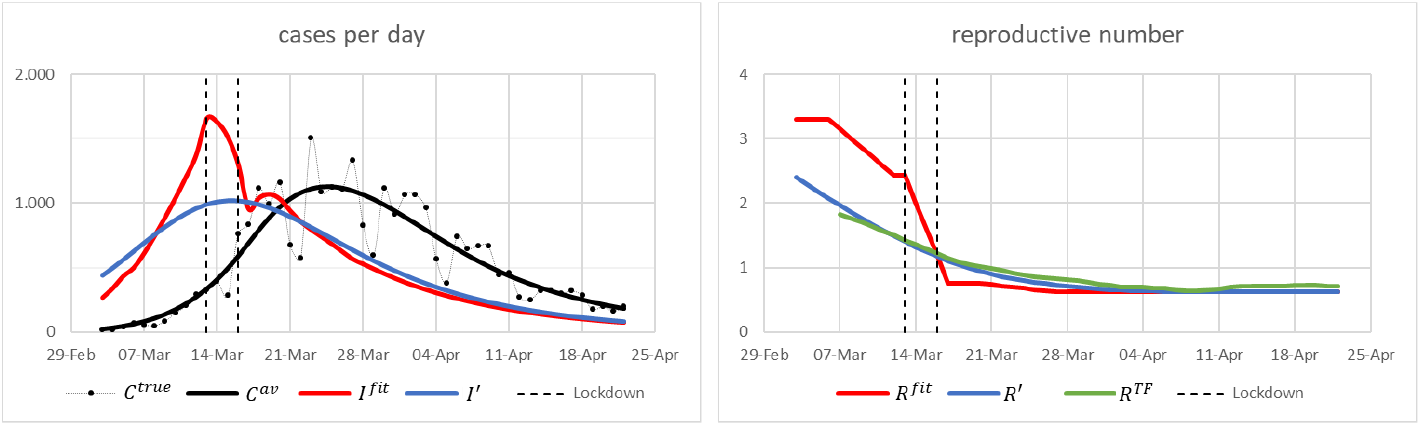
Cases and reproductive numbers in Switzerland around March 17. See text below for the definition of the curves.

**Figure 4:**
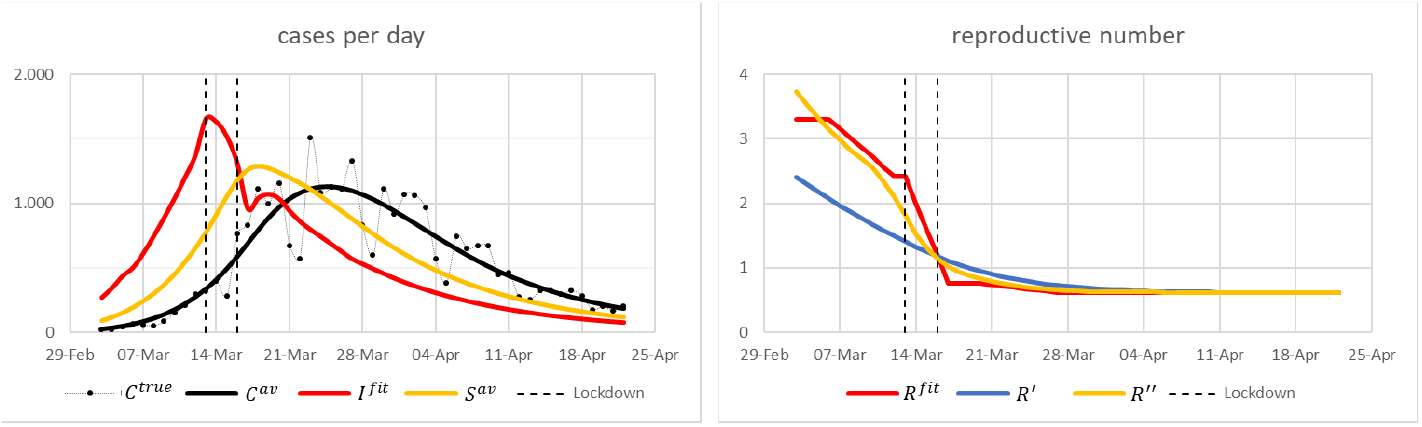
Left: ’True’ infection numbers (red) and their corresponding expected numbers of cases with symptom onset per day (yellow) and of confirmed cases per day (black). Right: The corresponding ’true’ reproductive numbers (red) and their reconstructions according to the schemes [7] (yellow) and [2](blue).

As in the main text, we denote by *w*_*n*_, *n* ≥ 1, the infection intensities and by *R*_*t*_ the effective reproductive number on day *t*.

We assume that no infections were present in Switzerland before *t*_0_ = February 24, 2020, i.e. for *t < t*_0_ we set *I*_*t*_ = 0.

The period between *t*_0_ and *t*_1_ = March 06 is taken as the burn-in phase, where we assume that each day *λ* infections are imported and that these already lead to secondary infections, i.e. for *t* = *t*_0_, *t*_0_ + 1, …, *t*_1_ − 1, we set

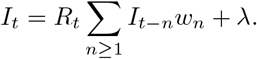

After *t*_1_, we assume that the ”internal” secondary infections dominate such that imported infections can be neglected, i.e. for *t* ≥ *t*_1_ the number of new infections *I*_*t*_ are given by

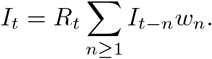

In particular, we assume throughout that the number of new infections is a deterministic function of the imported infections and of the reproductive numbers, i.e. that random fluctuations in the number of infections can be neglected.

A posteriori, these choices turn out to be consistent. According to our fit of the new infections per day, we get approximately 500 new cases for March 06. Thus, the neglected fluctuations are with a fairly large probability within twice the square root of 500, i.e. within *±*45, while the fitted value for *λ* is approximately 47.

We denote by *Z* the (random) number of days between infection and confirmation by virus test. Then the number of cases that are tested on day *t* and confirmed positive is given by

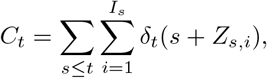

where *Z*_*s,i*_ are independent and identically distributed (i.i.d.) copies of *Z* and *δ*_*t*_(*x*) = 1, if *x* = *t*, and 0 otherwise.

We denote the probability measure of the common probability space of all these random variables by *P*, and expected values with respect to *P* by *E*. For the expected number 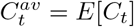 of confirmed cases on day *t* we get

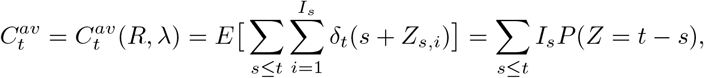

where we make explicit, that this series depends on the series of reproductive numbers, *R* = (*R*_*t*_), and on the number *λ* of daily imported infections during the burn-in phase.

Next, we denote by 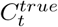 the true number of cases confirmed positive on day *t*. For determining the best fit, we use the *L*^2^-distance expression in the form^5^

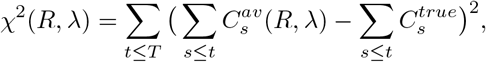

where the last information taken into account is that of *T* = April 27, 2020, the last number of confirmed cases not affected by the first relaxation of measures of April 27.

Then we set

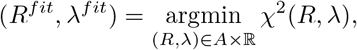

where *A* is a suitable set of admissible *R*-curves, encoding a priori assumptions to reduce the number of parameters to be fitted, which is necessary, because minimization over all *R*-curves would lead to overfitting. The a priori assumptions made by us are as follows.

It is reasonable to assume that the *R*_*t*_ do not fluctuate largely from day to day, except possibly around a limited number of ”change points”, where a change of measures is put in force. Therefore, we have chosen *A* to consist of piecewise linear *R*-curves. As we concentrate on the days around the lockdown, we limit ourselves to a minimal number of pieces. The natural choices for the change points are *t*_2_ = March 12 (the last day of ”normal life”), *t*_3_ = March 13 (the press conference announcing first drastic measures), *t*_4_ = March 17 (lockdown), *t*_5_ = March 20 (ban of gatherings *>* 5 people), *t*_6_ = March 27 (intermediate change point). After that, we assume that the reproductive number stays constant. (Here, the intermediate change point is introduced to test that after the ”ban of gatherings *>* 5 people” the reproductive number stays effectively approximately constant. If this was not the case, then the choice of change points needed revision.) Moreover, the case numbers are too low to estimate the reproductive numbers in the burn-in phase. Therefore, we rely for this period on typical estimates found in the literature and set *R*_*t*_ = 3.30, for *t* between February 24 and March 05. Finally, as up to March 20 measures are strengthened, but never relaxed, we require that the admissible *R*-curves do not increase before *t*_5_. Summarizing, we choose

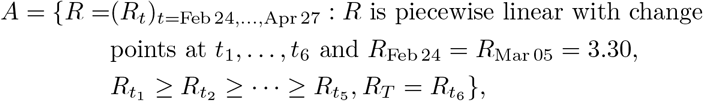

i.e. we fit 6 reproductive numbers, *R*_Mar 06_, *R*_Mar 12_, *R*_Mar 13_, *R*_Mar 17_, *R*_Mar 20_, *R*_Mar 27_, plus *λ*.

We would like to emphasize that the reproductive numbers resulting from this fitting procedure are quite unstable, in particular with respect to variations of the distributions of *Z* and of the infection intensities *w*_*n*_ that are not inferred here but estimated elsewhere [2, 8]. Therefore, *R*^*fit*^ should at most be considered a likely picture of reality. To get more confidence in the results, a thorough sensitivity analysis would be required, confidence intervals should be calculated (see e.g. the above mentioned [6]), and improvements to the analysis like the ones described in appendix B schould be incorporated.

## Appendix B Suggested improvements to the analysis

The simple analysis described above only gives a first indication of the reproductive numbers around the lockdown. In order to more precisely quantify the effects of the measures on the reproductive number, this analysis should be extended to include at least the following elements.

### Find change points in a systematic way

Instead of choosing the change points by a priori reasoning as above, one can infer them from the data by replacing the above *A* by

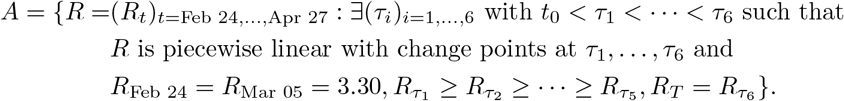

### Weekend effects

Weekend effects can be taken into account by substituting the above *C*_*t*_ by

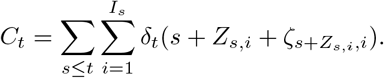

Here, *ζ*_*t,i*_, *i* ≥ 1, are i.i.d. copies of *ζ*_*wd*(*t*)_, where *wd*(*t*) *∈ {*Mon, Tues, …, Sun*}* denotes the weekday of *t. P* (*ζ*_Sat_ = 2) is then the probability, that a test that without weekend effects would have been performed on a Saturday was actually performed on the following Monday. These probabilities could be estimated directly from the confirmed cases, or they can be fitted together with the other parameters. For the expected number of confirmed cases on day *t* we get then

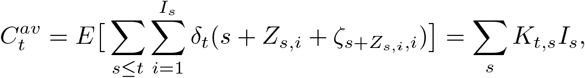

with

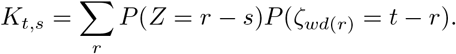

### Impact of quarantining on infection intensities

A natural guess for the infection intensities 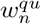 of those that quarantine starting the day after the onset of symptoms is

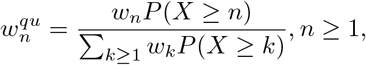

where *X* denotes the incubation time. The average infection intensities at day *t* are then

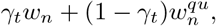

where *γ*_*t*_ *∈* [0, 1] is the proportion of infectives on day *t* that do not quarantine although symptomatic. Thus, quarantining can be taken into account by minimizing over *A×* ℝ *×G*, where *G* consists of an adequate set of admissible *γ*-curves.

## Acknowledgements

We thank Nicola Kistler for asking the right question and Erik Böttger, Jürg Fröhlich and Emanuel Wyler for helpful discussions.

## Footnotes

1 From a mathematical point of view, this is clear: If all the *I*_*t*_ are large, then we have *C≊ I ∗ p*, where *p*_*n*_ = *P* (*X* + *Y* = *n*), and 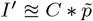, where 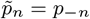, i.e. 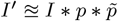.

2 See appendix A for the algorithm used to infer *I*^*fit*^.

3 We refer to [7] for the method applied to the cases with no known date of symptom onset.

4 As our focus is again on the days around March 17, we restrict our exposition to those days of infection where all infections can be assumed to be confirmed by the actual date of the monitoring. For a method to extend the reconstruction to later days we refer to [7].

5 The reason, why we choose the *L*^2^-distance between the cumulative counts and not between the daily counts, is that testing shows a weekly pattern due to non-corona related effects. Its impact shows less in the cumulative view. We refer to appendix B for a more direct way to take these effects into account.

